# A game theory based analysis of network connectivity reveals that state transition is crucial for epilepsy surgery outcome prediction

**DOI:** 10.1101/2021.11.19.21266569

**Authors:** Karla Ivankovic, Alessandro Principe, Justo Montoya, Linus Manubens-Gil, Mara Dierssen, Rodrigo Rocamora

## Abstract

Seizures recur in half of patients who undergo epilepsy surgery. Presurgical workup mainly focuses on seizures, but only partially helps predicting outcomes, even after invasive electroencephalography. We conceived a generalizable model to detect epileptogenic networks through connectivity changes and identified the crucial role of the transition from pre-seizure to seizure, achieving the highest outcome prediction (93%) to date in a chronological cohort of 21 patients with 3-year follow-up or more.

## Text

Epilepsy has a significant premature mortality burden up to nine times higher than that of unaffected population, especially when considering seizures and related accidents^1^. Twenty to forty million epileptic patients (0.25-0.5% of the world’s population, depending on prevalence estimates^2^) present uncontrollable seizures. Around half of these patients need an extended workup including invasive electroencephalography^3^. These studies provide data for modeling brain dynamics and connectivity to help diagnostics and treatment^4^, and are invaluable sources for neuroscientific modeling in humans^5^, thanks to their unmatched temporal resolution. Unfortunately, more than half of these patients recur at long term^6,7^. Understanding the activation of the epileptogenic network (EN) could become essential for treatment and fundamental for neuroscientific research. Here we describe a new model for predicting surgery outcome based on machine learning and game theory. Patients with a minimum 3-year follow-up were chronologically enrolled from our first stereotactic electrode placement until 2019. These criteria avoided as much as possible data selection that would add to other unavoidable biases, like seizure onset localization and electrode placement. The cohort included all lobar epilepsy types by subjects with good and poor outcome (see Supplementary Results). To assess whether our cohort would represent a solid ground truth and compare our results, a review of 1417 works was performed (Table 1).

**Table 1.** Literature review. **A comprehensive search of published literature until June 2021 was carried out with the following queries:** “*surgery outcome prediction AND epileptogenic zone”* (344 studies); “*epilepsy surgery AND postoperative outcome prediction”* (636), “*epileptogenic zone biomarker”* (113), “*epileptogenic zone AND epileptogenic network”* (324). Some parameters were evaluated only in those articles considering surgical outcome (SO), - = non-calculated.

| Parameters | SO% (works) | All |
| --- | --- | --- |
| Some *surgical outcome (SO)* assessment | 100% (21) | 1.50% |
| Post-surgical follow-up |  |  |
| <3 years | 95.2% (20) | - |
| ≥3 years | 4.8% (1) | - |
| SO classification (ROC-AUC) | 14.3% (3) | <0.01% |
| Data selection (by epilepsy type, seizure onset, other) | 90.5% (19) | - |
| Connectivity analysis | 61.2% (13) | - |
| Focus on seizure time | 52.4% (11) | 65% |

The majority of works, even existing models of seizure dynamics^8,9^, focus on seizure time; the rest rely on non-seizure time. Interestingly, we did not find studies focusing on pre-seizure time, usually considered for seizure forecasting^10^. To define the EN, we hypothesized that it is competing against non-epileptogenic networks (NN) to control their activity dynamics. The model produced network suggestions for comparison with actual surgical resections. Through functional connectivity analysis we were able to disentangle active from passive nodes by measuring changing connections (Figure 1). When seizures approximate, the EN’s changing connections should overcome the NN’s. To calculate the score of this hypothetical game and the connectivity change we assessed epochs using a support vector machine (SVM), node by node. Changing connections were measured by classification accuracies of paired epochs (considered as distinct classes), e.g., between non-seizure and seizure epochs. We used a game rule^11^ (maximin) to calculate gains across scenarios (random epochs) that represent possible game states, assuming that EN nodes would score better than NN nodes. We expected that far from seizures, EN nodes, which lack part of the regulatory feedback from NN^12^, would be more active but unable to turn the game balance (Figure 1), due to their limited number —the EN is smaller and surrounded by NN. During seizure time, conversely, both EN and NN should display significant connectivity change, the NN counteracting or falling under the influence of the EN (seizure propagation). However, during the pre-seizure time the EN activity should maximally differ from NN and therefore be distinguishable. We calculated how random network suggestions (noise) discern good from poor outcomes to assess the signal-to-noise ratio. Average noise was 62% because good to poor outcome ratio was higher than ½. Random score distribution reached high discrimination levels, and infinite iterations would eventually reach full discrimination. Still, their distribution has a limit, above which non-chance discrimination is found.

**Figure 1.**
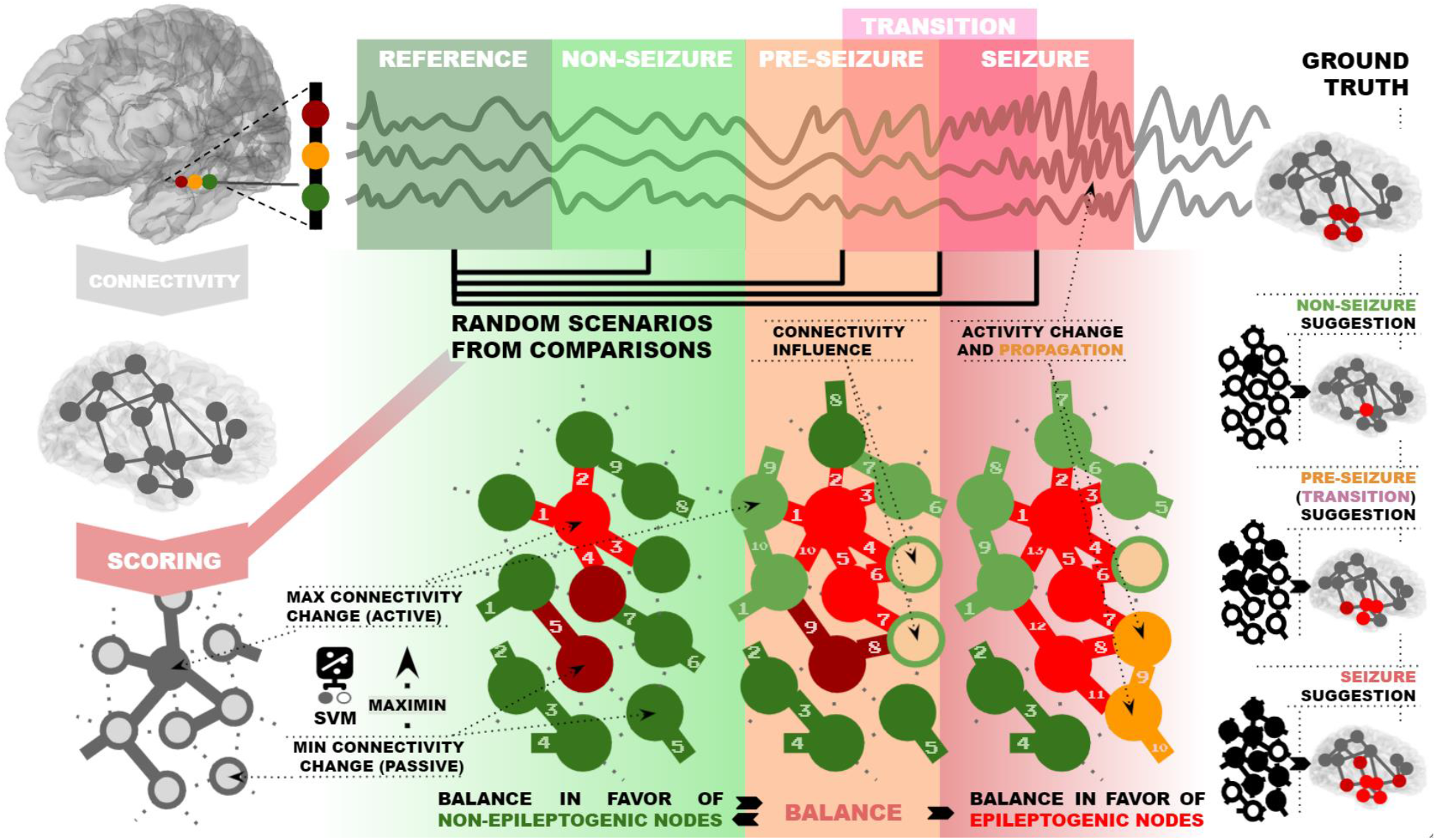
Model and methods summary. Thirty-two functional connectivity analyses and a SVM were used to assess connectivity change, reference were non-seizure epochs. The maximin rule scores nodes connectivity change. When either of networks sum up most of change (numbered connections) the balance is flipped.

As expected, connectivity wchange was low during interictal time, increased at pre-seizure, and burst during seizures (Figure 2A). However, it reduced to a minimum at seizure transition (the time covering equal proportions of pre-seizure and seizure). At this point of no return from progressing to seizure, most of the connectivity change is due to the EN. This unexpected but crucial finding shows that scoring the connectivity change can signal state transition. Indeed, outcome discrimination above noise limit was only possible at this time.

**Figure 2.**
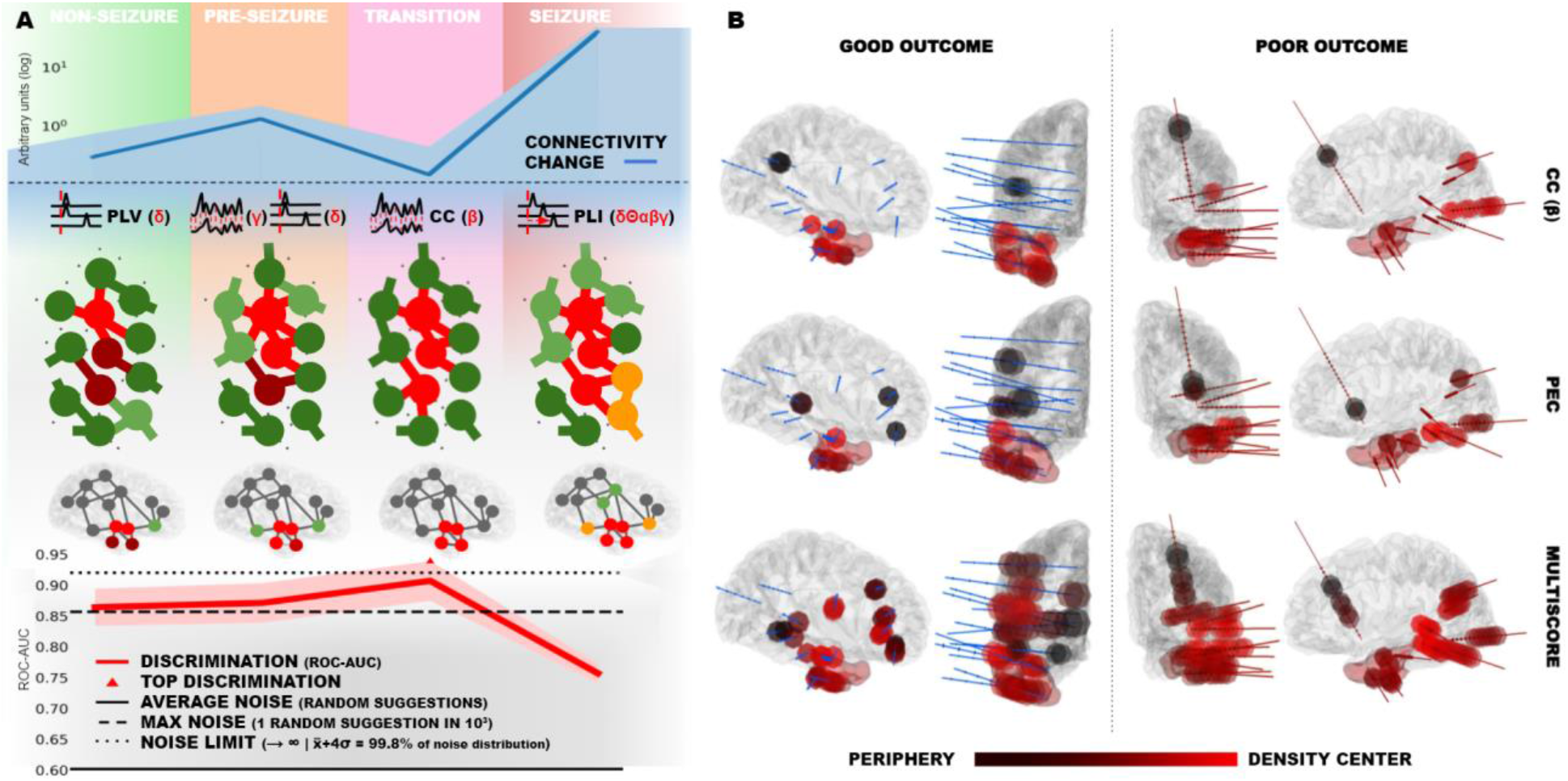
*A*. **Connectivity change and discrimination between outcome classes** as receiver-operator-characteristic areas-under-the-curve (ROC-AUC), both expressed as mean 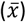 ± standard deviation (σ), along with best single-method models, graphically described and shortened as: PLV, phase-lock value; CC, cross-correlation and PLI, phase-lag index, Greek letters indicate EEG bands. *B*. **Scores applied to anatomy**. Most suggested EN nodes (dots) are within resection (red halo) in a good outcome patient; the opposite happens in a bad outcome patient. Lines represent multi-electrode positions.

Moreover, scores allowed an estimation of neural population activity that was extrapolated by comparing single-method with multi-method models (multiscores, see Methods). Far from seizures (epochs taken as far as possible from the seizure onset; Figure 2A; non-seizure), EN is represented by synchronized slow waves. At pre-seizure, slow and fast activity, which might represent NN nodes counteracting the EN, distinguish the EN from the NN. At transition, slow and fast activity converge to mid-range waves, indicating that the EN maximizes its ability to influence the NN. Finally, during seizures, outcome discrimination is poor and achieved through all EEG bands, which could mark specific activities of NN nodes that react or fall under EN influence. Notably, the most consistent functional connectivity method was prediction error connectivity^13^ (PEC), a non-frequential method that emphasizes temporal relations between signals. This fact is in line with the central concept of the model, which allows spatial estimation of the network at specific times, especially at state transitions. This concept could be successfully applied to describe neural network states during neuropsychological events, pinpointed by connectivity change minima.

Furthermore, future versions of our algorithm could have clinical applications (Figure 2B) helping clinicians design surgeries, as single-method scores, and indicate surgery feasibility, as multiscores. As shown in the example (Figure 2B), several multiscore nodes are close to eloquent areas in the poor outcome patient. The network game concept might also contribute to seizure forecasting and solve controversies on epilepsy network theory^14^. Indeed, future studies will define whether the EN is constantly striving to enter the pre-seizure state, which is the gateway to transition. Moreover, further insight is needed to understand the mechanisms of connectivity change and the timing of transition, which were only partially explored in this work. These studies will allow us to generate reliable brain connectivity models based on solid ground truth.

## METHODS

### Revision of the literature

To evaluate the current state of the art, we screened the publications focused on epileptogenic network (EN) localization and surgery outcome prediction, published before June 2021.

#### Database

A systematic search was conducted in the PubMed database (National Institutes of Health (NIH) which has the major digital archive of papers in the biomedical and life sciences.

#### Keywords

The PubMed database was searched using four sets of keywords (see Table 1). This review covers only the literature on epileptogenic network (EN) localization and surgery outcome prediction. The terms “surgery outcome prediction”, “epilepsy surgery”, “epileptogenic zone”, collected into MeSH (Medical Subject Headings), were used as primary keywords. After reading 10% of the papers, the secondary searches were defined as: “surgery outcome prediction AND epileptogenic zone”, “epilepsy surgery AND postoperative outcome prediction”, “epileptogenic zone biomarker”, “epileptogenic zone AND epileptogenic network”.

#### Inclusion criteria

The period investigated was until June 2021 and only papers written in English were included. The papers selection was performed by considering the keywords that appeared in the title and abstract. Then each paper was selected for its content. Studies that focused on EN localization and surgery outcome prediction were used.

#### Exclusion criteria

Doctoral theses, case reports, reviews and expert opinions were excluded from this investigation.

#### Parameters

All search results were categorized as focusing either on non-seizure (connectivity far from seizures, resting state, sleep or isolated interictal spikes), on pre-seizure (specified time frame preceding seizure onset) or on seizure time. If a study analyzed multiple time frames including seizures, we considered it to be seizure-focused, since local-field potential dynamics and connectivity changes are maximal during seizure time, as demonstrated by our results. Studies analyzing magnetic resonance imaging, magnetoencephalography, or positron emission tomography, as compared to the seizure onset zone defined by EEG, were categorized as seizure focused. Next, we checked whether studies distinguished between surgery outcomes and restricted our analysis to publications reporting some surgery outcome classification. Since few works reported surgery outcome classification by receiver operating characteristic-area under the curve (ROC-AUC), any type of quantified or qualitative comparison between outcomes was considered (Z-score, P-value, or qualitative comparison). Only studies based on invasive EEG data analysis (SEEG, subdural strips or grids) were included. The selected studies were further categorized by data selection, epilepsy type, patient cohort size, recording type, surgery outcome comparison method, post-surgical follow-up duration, analytical strategy, and connectivity assessment (Supplementary Table 1). The most important discriminants considered were data selection and surgical outcome classification or description based on resection. Studies were tagged with data selection whenever patients were selected by epilepsy type, lesion type (e.g., by focal cortical dysplasia), seizure onset pattern, suboptimal ictal recording, clinical onset preceding the seizure onset, or lack of information. Works were labeled to use true classification only when ROC-AUC was employed to assess the surgical outcome. The connectivity-based analytical strategies considered were: cortico-cortical evoked potentials^15^, functional connectivity by nonlinear regression^16^, mutual information^17^, Spearman correlation^18^, Granger causality^19^, graph eigenvector centrality^20^, betweenness centrality and computational network models. Approaches to local field-potential (LFP) analysis, not accounting for connectivity, were considered the ones focused on seizure onset patterns^21^, high frequency oscillations (HFOs)^22^ including high gamma oscillations, preictal spikes and ripples, or single-channel low frequency phase-high frequency amplitude coupling^23^. We set apart studies where outcome description or classification was obtained without direct reference to surgical resection, but to an alternative reference based on signal analysis (e.g., epileptogenicity index^24^) or subjective estimation of the seizure onset zone, even when performed by trained epileptologists.

### Patients and SEEG recordings

SEEG data recorded from a retrospective cohort of drug-resistant epileptic patients chronologically admitted to the Epilepsy Unit of Hospital del Mar from 2013 to 2018 were analyzed. The only inclusion criteria were a postsurgical follow-up of a minimum of 3 years and a focal epilepsy treatable with non-palliative surgery. Vagal nerve stimulation or other device implantation and callosotomy were considered as palliative surgical approaches. Patients without any spontaneous seizures recorded during SEEG were also excluded. All recordings were performed using a standard clinical EEG system (XLTEK, subsidiary of Natus Medical). Implantations were performed using intracranial electrodes (Dixi Médical, Besançon, France; diameter: 0.8 mm; 5 to 15 contacts, 2 mm long, 1.5 mm apart), which were stereotactically inserted using robotic guidance (ROSA, Medtech Surgical, Inc). The implantation depended entirely on clinical criteria, without reference to this study. SEEG data of a single, randomly chosen seizure were extracted for each patient. The extracted data included a period preceding the seizure of at least 5 minutes and the whole seizure to account for the considered times: pre-seizure, transition from pre-seizure to seizure, and seizure. For the analysis of non-seizure time, the data was extracted as far as possible from the preceding seizure or recording start, and 24h after electrical stimulation. Data was extracted in EDF+ format as bipolar channels, and the metadata included seizure onset and end annotations, channel labels and the sampling frequency. Only SEEG recording channels were used, scalp EEG and other types of recordings were excluded.

### Analytical framework

The framework for EN identification from SEEG data was implemented in Python (version 3.7.10), using the modules: *numpy, scipy, matplotlib, itertools, collections, pickle*, and *pyedflib*. Modules for data management (*core, data_legacy* modules) and processing (*eeg* module) were developed in-house. The final pipeline was implemented for preprocessing and analysis. At pipeline initialization, epoch span was set to 1 second, the overlap step to 500 ms, and the events of interest were defined. Epochs were defined by overlapping 1-second windows and a specific number of random data splits was considered. These parameters were not further explored and remain to be possibly tuned. Time frames were defined in a custom fashion relative to seizure length, with a working assumption that a longer pre-seizure phase would precede a longer seizure. Further exploration of the time domain and use of fixed time windows for all seizures might provide further insight on the features of pre-seizure and transition time.

#### First-stage preprocessing

Raw data in the *EDF+* format, containing SEEG signal and metadata, were loaded as a table. A mask overlay was constructed to label the signal time-domain epochs per event type (non-seizure, pre-seizure/transition, and seizure), relative to the clinical annotations of seizure start and end (Supplementary figure 1A). The position of the mask was optimized so that epochs’ start fitted annotations with minimal delay. All epochs were initially tagged as non-seizure. Depending on the analysis, time frames of interest were defined relative to seizure length (*L*). To analyze *pre-seizure*, epochs were tagged as pre-seizure when within the interval preceding 0.6·*L* the seizure onset. To analyze the *transition from pre-seizure to seizure*, epochs were tagged as pre-seizure to seizure transition when within the interval spanning from 0.3·*L* before to 0.3·*L* after seizure onset. To analyze *seizure*, epochs between seizure start and end were tagged as seizure. To analyze the baseline connectivity change, non-seizure data were split in half and the halves were separately tagged. Thirty random epochs of each event (overall 60 epochs) were then selected and combined for further preprocessing.

#### Second-stage preprocessing

Most but not all recordings were sampled at 500/512Hz, thus some recordings had to be homogenized: signals sampled at 250 Hz were up-sampled to 500 Hz, while signals sampled at 1024 Hz and 2048 Hz were down-sampled to 512 Hz. Power frequency noise (50 Hz) was removed by a notch filter. To account for different connectivity quantification approaches, we implemented frequency domain connectivity analyses, including low frequency phase-high frequency amplitude coupling between node pairs (PAC), phase lag index (PLI), phase locking value (PLV), real part of the spectral coherence (SCr) and imaginary part of the spectral coherence (SCi), and time domain connectivity analyses, including cross correlation (CC) and prediction error connectivity (PEC). PAC is commonly analyzed in a single node, however, a custom PAC analysis was implemented to compute phase locking between low frequency phase of one node and high frequency amplitude phase of another node, to account for network connectivity. For PLI, PLV, SCr, SCi and CC analysis, the signal was filtered to specific frequency bands: δ (1-4Hz), θ (4-8Hz), α (8-12Hz), β (13-30Hz), low γ (30-70Hz), high γ (70-150Hz). Low frequencies in the δ range and high frequencies in the low γ range were considered for PAC analysis. PEC was applied on the whole-band signal. Connectivity analysis with each of 32 connectivity methods (five methods in six different frequency bands, PAC and PEC) was applied to all 60 epochs, yielding connectivity matrices with dimensions N·N·60, where N is the number of nodes (Supplementary figure 1A). Connectivity matrices were saved as a custom-format prep file.

#### Analysis

For each node pair, all vectors containing epoch connectivity values were selected from the connectivity matrix. Vectors were passed to the support vector machine (SVM) classifier as features, whereas the epoch tags were the target (Supplementary figure 1A). The radial basis function was used as the kernel for building the SVM model. Data were randomly split into training and testing sets using the K-fold method with *k*=6 folds, thus approximately one fold every 10 seconds. In our model, EN and NN nodes are agents in a hypothetical game, where each agent can change the connectivity with other agents in several ways (see Main, Figure 1). These possibilities of connectivity change are modelled using the K-fold random data splits, so a fold represents a possible connectivity evolution. Since connectivity at a specific time point could be the result of an infinite array of branching points in the past, tracking the connectivity evolution should consider all possible comparisons between epochs, and eventually sub-epochs. Branching points should display maximal connectivity changes when compared with final connectivity. Since the analysis of all possible branching points would require powerful cluster computing, and therefore would limit the applicability of the framework, random epoch comparison was used instead. Thus, randomly selected points, considered as generators of chains of connectivity events up to random current epochs, were analyzed. These chains of events were dubbed game scenarios. The outcome of a game scenario was scored by a cross validation score (CVS), which was computed for each fold. The nodes with prominent connectivity changes have a higher probability of achieving a high CVS in a random fold, in comparison to the nodes with less important connectivity changes. To evaluate the probability of the connectivity change in the nodes, the maximin function (a decision rule used in game theory for maximizing the minimal possibility of winning) of the CVSs was computed. The node pairs were then sorted by the maximin values. Since the game outcome is seizure, the top-scoring nodes were considered epileptogenic. The classification was then repeated, this time starting with the vectors of the highest-scoring node pairs, to which other node pairs could further contribute. If the new score was higher or equal to the previous best score, the classification was repeated. The iteration continued until the maximin score decreased. In other words, putative EN nodes were added up until new nodes could not contribute anymore to the discrimination power between epoch classes. The top-scoring nodes were selected as the EN, CVSs and maximin scores were saved as a custom *res* file format.

**Supplementary Figure 1.**
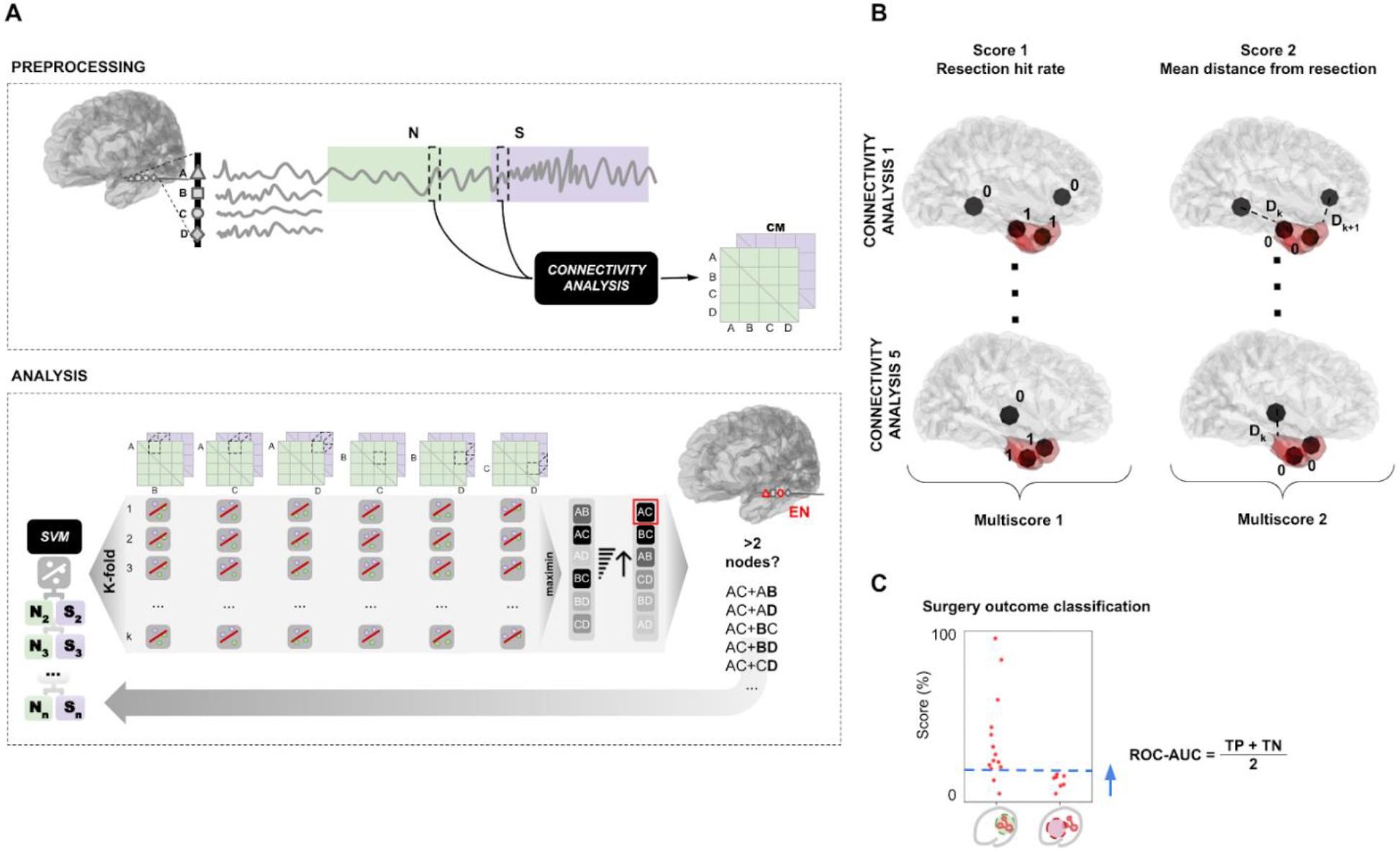
Schematic of the computational workflow. *A*. **Schematic of the model implementation for EN definition**. Signal epochs of two classes (N - reference (green), S - event (purple)) were processed across node (A, B, C, D) pairs by a connectivity analysis. Individual vectors (dashed cubes) of the connectivity matrix (CM) were used as features for machine learning classification by the support vector machine (SVM). The K-fold, random data-splitting method, was used for simulating scenarios of connectivity change. The maximin function was used to score the connectivity change of node pairs. The node pairs were sorted (upward arrow) by their maximin score, and the top-scoring nodes (red square) were selected as EN (red nodes). The classification was repeated (grey arrow), adding the next top-scoring node pair, until the maximin score decreased. *B*. **Schematic representing computation of scores**. Multiscores were calculated by averaging Scores 1 and 2, across separate connectivity analyses, up to combinations of five. *C*. **Schematic representing good and poor surgical outcome classification by a moving threshold** (blue dashed line). ROC-AUC is computed using Score1, Score2, Multiscore 1 or Multiscore 2 as a predictive measure. A correct surgical resection is represented by a green dashed circle of the epileptogenic network (red nodes and edges), and an incorrect resection by a red dashed circle.

### Connectivity change computation

The analytical framework selects a network of nodes with dominant connectivity changes. What if all or almost all implanted nodes are finally selected? In this case, the predicted network likely contains non-epileptogenic nodes (Figure 1), making the distinction impossible. Thus, we considered the degree of connectivity change for the analyzed time intervals (non-seizure, pre-seizure, transition and seizure) by calculating the number of networks overcoming a size threshold. The threshold was network-specific, accounting for variation between connectivity analyses and patient implantation schemes. For each connectivity analysis, network size range across patients was analyzed. For each patient, network size (*NEN*) was normalized by the overall number of implanted nodes (*NI*), expressed as a ratio (*R*):

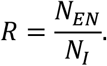

The final upper size threshold (*T*) was set as four standard deviations (σ) above average network size:

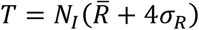

which is a statistical limit including >99.8% of data. Setting the lower size limit was unnecessary since the analysis algorithm controls for the selection of a minimal number of nodes. The mean and standard deviation of the number of networks exceeding the threshold across connectivity analyses was computed for each time interval.

### Reality check: comparison between network suggestions and surgical resections

Brain volume reconstructions for individual patients were made from pre- and postoperative CT and MRI scans. Resection volumes were inferred from the postoperative reconstructions. 3D Slicer (version 4.10.2), an open-source software for image analysis and scientific visualization with Python implementation, was used^25^. Each reconstruction included precise coordinates of implanted electrodes. New modules were coded to visualize network suggestions within the 3D Slicer environment. The predicted EN was interposed to the virtual scene by highlighting the electrode contacts with matching labels. Network topology was evaluated relative to the original resection by computing scores (Supplementary figure 1B). Position of each electrode of the bipolar source was analyzed relative to the resection volume. Two scores were calculated to compare network suggestions with surgical resections. Score 1 was expressed as the ratio of electrodes that fell within the resection:

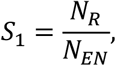

where N_R_ are the suggested nodes within the resection boundaries and N_EN_ are all suggested nodes. An alternative score, S_2_, was computed using Euclidean distances from the electrode center to the closest resection surface. If the node was within resection, the distance was set to zero. The alternative score was expressed as the mean distance from resection of the suggested nodes.

### Multiscores

To combine different connectivity aspects for network definition, multi-method scores (multiscores) were computed by averaging single-method scores (S_1_ or S_2_; Supplementary figure 1B). Groups of connectivity methods were formed, with all possible combinations from two to five methods. Scores obtained by separate connectivity methods were averaged across all possible combinations:

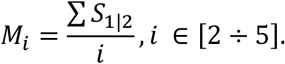

### Surgery outcome classification

The surgery outcome was evaluated relative to the Engel scale, with Engel IA-D classified as good and Engel ≥ II classified as poor outcome. Hypothetical ENs were identified in each patient using 32 connectivity analyses. Suggested networks exceeding *T* (see Connectivity change computation paragraph) were assigned the worst possible score depending on the patient outcome. Single scores and multiscores were used to classify surgery outcomes. The scores were sorted, and a moving threshold was applied from 0 to 100% of the highest score, with a step of 0.1% (Supplementary figure 1C). The optimal threshold to identify surgery outcome based on the score was not known *a priori*. Good and poor surgery outcomes were considered as true positive (TP) and true negative (TN), respectively. ROC-AUC was calculated at each threshold, as:

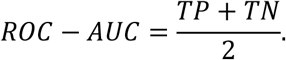

### Chance level distribution

To evaluate the degree of network discrimination, signal-to-noise ratio was assessed by testing the surgery outcome classification based on random network suggestions, which consisted of random node sets, ranging from 1 to all possible nodes. Chance ROC-AUC for surgery outcome classification was computed using S_1_. The test was iterated 1000 times. As the maximum ROC-AUC may change with iterations, eventually reaching perfect discrimination after near infinite iterations, the noise limit was set to four standard deviations above average chance ROC-AUC, which accounts for more than 99.8% of random scores.

### Connectivity method ranking through network comparison

Method informativeness in EN definition was evaluated by comparing single-method networks to the union of networks that yielded high multiscores. The smallest size of the groups of methods prompting the best ROC-AUC for a time frame was considered, as fewer methods should be more informative than more methods with equal ROC-AUC value. Combinations with above-average ROC-AUC, for the respective group size, were selected. Each single-method network (N) was then compared to the network union (U) of methods comprising a combination of size *i*, from 2 to 5. The network overlap ratio (*RO*) was calculated by dividing the number of shared nodes by the number of nodes in *U*:

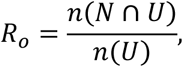

where *n* is a function mapped to the number of elements in a set. The average *RO* was calculated for each single-method.

### Network topology colormap

Electrodes comprising the selected network nodes were considered. The putative center of the network in three-dimensional space was computed as a point cloud centroid, by averaging coordinates of all electrodes in each dimension (x, y, z) separately. The Euclidean distances from the center to each electrode were scaled from 0 to 1. The electrodes were labelled with an aura colored relative to the respective distance value.

### Statistical analysis

Python libraries Numpy^26^ (version 1.20.0), Scipy^27^ (version 1.6.3) and Matplotlib^28^ (version 2.1.0) were used for statistics and graphics.

## Supporting information

Supplemental Results

## Data Availability

All data produced in the present study are available upon reasonable request to the authors.

## CODE AVAILABILITY

The code repository for the analysis framework is available at https://github.com/ivkarla/epigame-y1.

