## Supplemental Results for "A game theory based analysis of network connectivity reveals that state transition is crucial for epilepsy surgery outcome prediction"

3 Epilepsy Unit - Neurology dept. Hospital del Mar - Parc de Salut Mar - Barcelona, Spain. Member of the European Reference Network EpiCARE.

4 Bio-engineering faculty, internship fellow – University Pompeu Fabra, Barcelona, Spain

5 SEU-Allen Joint Center for Neuron Morphology, Southeast University (SEU), Nanjing, China

6 Centre for Genomic Regulation (CRG), The Barcelona Institute of Science and Technology, Barcelona, Spain

7 Biomedical Research Networking Center on Rare Diseases (CIBERER), Institute of Health Carlos III, Madrid, Spain

* These authors equally contributed to this work.



**Abstract**

Seizures recur in half of patients who undergo epilepsy surgery. Presurgical workup mainly focuses on seizures, but only partially helps predicting outcomes, even after invasive electroencephalography. We conceived a generalizable model to detect epileptogenic networks through connectivity changes and identified the crucial role of the transition from pre-seizure to seizure, achieving the highest outcome prediction (93%) to date in a chronological cohort of 21 patients with 3-year follow-up or more.

### SUPPLEMENTARY RESULTS

#### Meta-analysis

Selected studies were all retrospective. Only two of 21 studies maximally avoided data selection, such as patient selection due to their epilepsy type, seizure-onset pattern, or diagnostic certainty. A ratio of studies focused only on temporal lobe epilepsy (TLE) was calculated. TLE is indeed the most common type of epilepsy, but also the lobar epilepsy with the best surgical outcome rate. The median cohort size was 18 patients (Q1=16, Q3=34). Patient follow-up varied widely from 3 months to 20 years or more. However, only one study considered a minimum follow-up of at least 3 years. True surgery outcome classification —specificity and sensitivity for distinguishing two or more outcome classes— was performed only in 3 studies, corresponding to 0.002% of all retrieved scientific papers (Table 1). Importantly, not all works used the surgical resection as their main reference when describing or computing outcome classification. Several studies relied on expert opinion, which is the visual detection of the seizure onset zone, or other subjective criteria. However, resection was the main reference for most studies (15 of 21, 71.4%). As expected from the current view of the epileptogenic zone as a network, connectivity analysis overcame the separate analysis of brain region LFPs. Accordingly, it was more likely that a connectivity study would quantify the surgery outcome classification by ROC-AUC, as only 1 out of 9 LFP-based studies performed such quantification. A ROC-AUC of 87.5% was the best reported classification, in a work where a smaller cohort without clear temporal bounds and an incomplete lobar epilepsy representation was considered (Supplementary Table 1). A long array of methods and theories have been suggested to explain seizure dynamics and their occurrence, but only a handful of them are based on a solid ground truth, which could foster the development of clinical applications. For this reason, this study was conducted as similarly as possible to a clinical trial.

#### Patient epilepsy, outcome, and follow-up

Among an overall cohort of 50, 21 patients were chronologically enrolled. The cohort size was above the median size for similar studies (see Meta-analysis). All kinds of lobar epilepsy were present in our dataset, including occipital and insular lobe epilepsy, which are more difficult to study with other iEEG techniques. 14 patients had good surgery outcomes, while seven had poor surgery outcomes (Supplementary Table 2). Sampling rates differed between patients, with one patient recorded at 250 Hz, 16 patients at 500 Hz, two patients at 512 Hz, one patient at 1024 Hz and one at 2048 Hz. Length of the seizures ranged from 14 s to 505.5 s (mean 119.3 s; median 91.2 s).

Supplementary Table 1. **Meta-analysis of 1417 publications in the PubMed database revealed 21 studies reporting surgery outcome comparison based on iEEG analysis.** The studies are grouped by keyword sets.

| Paper | Strategy | Uses connectivity | AUC | Recording type | Cohort size | Minimal follow-up (years) | Epilepsy | Resection as ground truth | Data selection |
|---|---|---|---|---|---|---|---|---|---|
| Keywords: surgery outcome prediction AND epileptogenic zone | | | | | | | | | |
| Guo et al. 2020 Clin. Neurophysiol. | Cortico-cortical evoked potentials | Y | N | SEEG | 25 | 1 | F, I, O, P, T | N | Y |
| Lagarde et al. 2018 Brain | Nonlinear regression | Y | N | SEEG | 59 | 3 | F, O, P, T | N | Y |
| Lagarde et al. 2018 Epilepsia | Seizure onset patterns | N | N | SEEG | 143 | 1 | F, O, P, T | N | Y |
| Rummel et al. 2015 PLoS ONE | LFP and mutual information | Y | N | iEEG | 16 | 1 | F, P, T | Y | Y |
| Tomlinson et al. 2017 Epilepsia | Synchronicity (Spearman correlation) | Y | N | iEEG | 17 | 2 | F, P, T | N | Y |
| Hussain et al. 2017 Epilep. Res. | HFO | N | N | iEEG | 60 | 2 | T, extra-T (U) | Y | N |
| Tamilia et al. 2021 Ann. Neurol. | Ripple propagation | N | N | Surface EEG, iEEG, MEG | 28 | 1 | F, O, P, T | Y | Y |
| Keywords: epilepsy surgery AND post-operative outcome prediction | | | | | | | | | |
| Goodfellow et al. 2016 Sci. Rep. | Neural mass model | Y | 87.5% | iEEG | 16 | 2 | F, P, T | Y | U |
| Park et al. 2012 Clin. Neurophysiol. | HFO | N | N | iEEG | 16 | 1 | F, P, T | Y | Y |
| Kuroda et al. 2021 Brain Comm. | HFO+phase-amplitude coupling | N | 82% | iEEG | 135 | 1 | T, extra-T (U) | Y | Y |
| Keywords: epileptogenic zone biomarker | | | | | | | | | |
| Li et al. 2019 EMBC | preictal spikes, fast activity, low frequency suppression | N | N | SEEG | 24 | 2 | F, O, P, T | Y | Y |
| Qi et al. 2020 Front. Hum. Neurosci. | LFP | N | N | SEEG | 19 | 1 | F, I, T | Y | Y |
| Ren et al. 2019 J Neurol. | Granger causality and graph analysis | Y | | iEEG | 7 | 2 | T | Y | Y |
| Brázdil et al. 2017 | LFP | N | N | SEEG | 10 | 1 | T, extra-T (U) | Y | Y |
| Bandarabadi et al. 2019 Ann. Neurol. | Phase locking and HFO | Y | N | SEEG | 16 | 1 | U | Y | Y |
| Grobelny et al. 2018 Clin. Neurophysiol. | Graph analysis | Y | N | SEEG | 36 | 1 | T | Y | Y |
| Tamilia et al. 2018. Ann. Neurol. | Ripple propagation | N | N | iEEG | 27 | 1 | F, I, O, P, T | Y | U |
| Tomlinson et al. 2016 Front. Neurol. | Interictal propagation patterns | Y | N | iEEG | 18 | 2 | F, T | N | N |

| | | | | | | | | | |
|---|---|---|---|---|---|---|---|---|---|
| Keywords: epileptogenic zone AND epileptogenic network | | | | | | | | | |
| Sinha et al. 2017 Brain | Dynamical network model | Y | 81.3% | iEEG | 16 | 1 | F, O, P, T | Y | Y |
| Li et al. 2018 Network Neuroscience | Graph analysis | Y | N | SEEG, iEEG | 42 | 1 | F, O, T | N | Y |
| Sip et al. 2021 PLoS Comput. Biol. | Computational model of seizure recruitments and propagation | Y | N | SEEG | 18 | 1 | F, I, P, T | Y | Y |

HFO – high frequency oscillations; iEEG - invasive EEG; LFP – local field potential; MEG - magnetoencephalography; Y - yes; N - no; F - frontal; I - insular; P - parietal; O - occipital; T - temporal; U - unspecified

Supplementary Table 2. **Patients' demographics and clinical facts**

| Characteristic | Total | Good outcome (Engel I) | Poor outcome (Engel ≥ II) |
|---|---|---|---|
| n (F) | 21 (12) | 14 (7) | 7 (6) |
| Age at surgery (median) | 22 - 57 | 22 - 57 (41) | 22 - 46 (32) |
| Years of epilepsy at surgery (median) | 5 - 46 | 8 - 46 (23) | 5 - 33 (14) |
| Average follow-up in years (σ) | 5 (2) | 6 (2) | 5 (2) |
| Left/right hemisphere | 12/9 | 7/7 | 5/2 |
| With/without focal MRI findings | 13/8 | 7/7 | 6/1 |
| N electrodes (median) | 5 - 19 | 5 - 17 (11) | 8 - 19 (14) |
| N contacts (median) | 44 - 217 | 44 - 185 (123) | 113 - 217 (142) |
| Epilepsy | | | |
| Temporal mesial | 11 | 9 | 2 |
| Temporal neocortical | 2 | 1 | 1 |
| Insular | 1 | 0 | 1 |
| Frontal | 2 | 1 | 1 |
| Parietal | 4 | 2 | 2 |
| Occipital | 2 | 1 | 1 |

F-female; n-number of patients; N, number of electrodes or contacts; σ, standard deviation.

### Network scores and surgery outcome prediction

$S_2$ yielded inferior accuracy of surgery outcome prediction (maximum 89%, using multiscores at pre-seizure to seizure transition), therefore we focused on $S_1$ as a predictive measure. Single-method scores allowed maximal surgery outcome prediction ROC-AUC of 82% during non-seizure, pre-seizure and transition time (median 68%, 68% and 71%, respectively), and 71% during seizure time (median 57%;

Supplementary figure 2). Multiscores increased the accuracy of outcome prediction. The number of multiscore combinations increased exponentially with group size, reaching 201,377 combinations using five methods. We stopped the iteration at the combinations of five as the ROC-AUC distributions were stable across combinations of three, four and five (Supplementary figure 2), and the top scores were yielded by <0.001% of the combinations. Baseline connectivity change, computed during non-seizure time, allowed a maximal surgery outcome prediction performance of 89% (mean 70%) only using multiscore combinations of five connectivity methods. Pre-seizure time achieved a maximal ROC-AUC of 89% (mean 70%) with several multiscore combinations, from three to five. At transition from pre-seizure to seizure a maximum of 93% (mean 72%) was reached with four-method multiscore combinations. A maximum of 79% (mean 55%) was obtained by four-method combinations at seizure. Interestingly, the interquartile range decreased with combination size only using seizure time. Surgery outcome prediction was further assessed by computing noise distribution obtained by random network suggestions. The average noise was 60%, with the standard deviation of 8%. The average value was higher than the general chance level (50%) likely due to the disbalance between class sizes (*good:poor*=2:1).

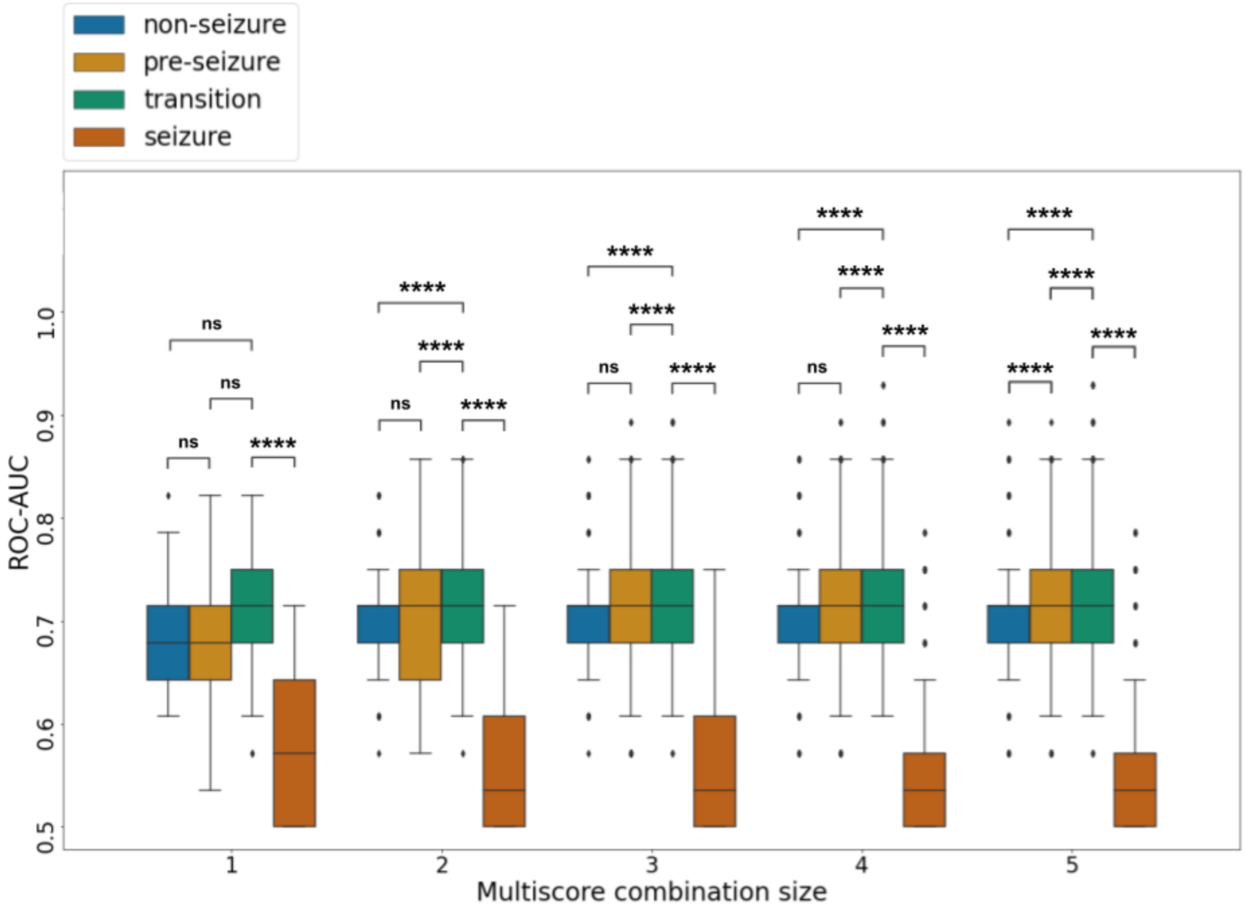


Supplementary Figure 2. **ROC-AUC distributions achieved by single Scores 1 obtained using 32 connectivity methods (1) and by multiscores of all possible combinations of two to five methods (2-5)**.
The highest discrimination of 93% was achieved by combinations of four methods in transition time frame (green). P-value annotation: ****: P <= 0.0001; ns: non-significant, Mann–Whitney U two-sided test with Bonferroni correction. Data represent grouped data per time frame, including the group median (line), interquartile range (IQR; box), span of 1.5·IQR past the low and high quartiles (~99.9% of data; whiskers), and the outliers (diamond symbol).

**Connectivity method ranking through network comparison**

Single-method scores and therefore connectivity analyses were ranked by comparing associated network suggestions (node sets) with multiscore suggestions, which provided better class discrimination. Ranking methods provided us insight on the possible mechanisms governing the connectivity changes at different time frames (see main text and figure 2). Supplementary figure 3 lists all methods per time frame.

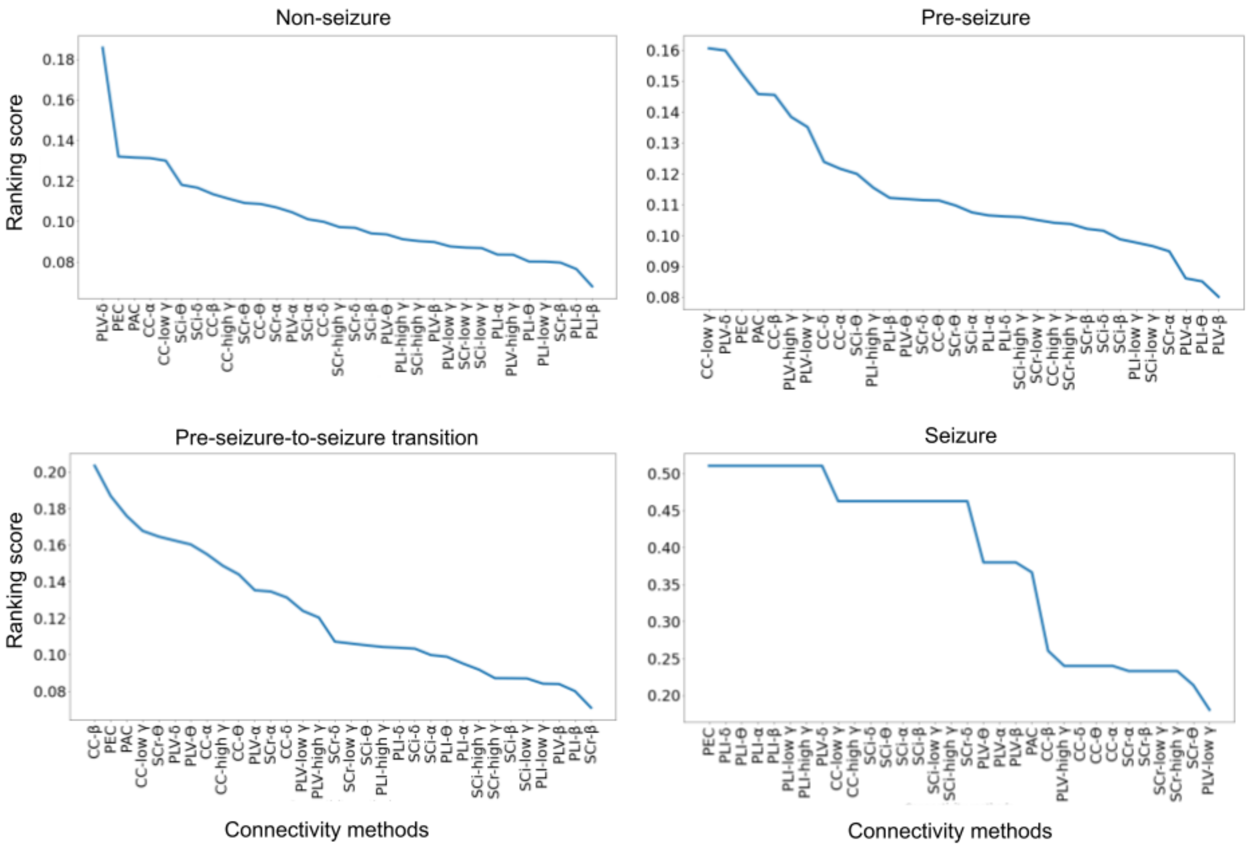


Supplementary Figure 3. **Connectivity methods and respective frequency bands by their informativeness in epileptogenic network definition, in different timeframes.**

Ranking score units are arbitrary and represent the overlap of the single-method network with the union of networks associated to multiscores above average ROC-AUC values (non-seizure 70%; pre-seizure 70%; transition 72%; seizure 55%). CC - cross correlation; PAC - low frequency phase-high frequency amplitude coupling; PEC - prediction error connectivity; PLI - phase lag index; PLV - phase locking value; SCi - imaginary part of the spectral coherence; SCr - real part of the spectral coherence.
